# Predicting individual risk for COVID19 complications using EMR data

**DOI:** 10.1101/2020.06.03.20121574

**Authors:** Yaron Kinar, Alon Lanyado, Avi Shoshan, Rachel Yesharim, Tamar Domany, Varda Shalev, Gabriel Chodick

**Author notes:** **Corresponding author** Prof. Gabriel Chodick, PhD Maccabi Institute for Research & Innovation, Israel Keufman St. 4, Tel Aviv Israel 68125.

## Abstract

**Background:** The global pandemic of COVID-19 has challenged healthcare organizations and caused numerous deaths and hospitalizations worldwide. The need for data-based decision support tools for many aspects of controlling and treating the disease is evident but has been hampered by the scarcity of real-world reliable data. Here we describe two approaches: a. the use of an existing EMR-based model for predicting complications due to influenza combined with available epidemiological data to create a model that identifies individuals at high risk to develop complications due to COVID-19 and b. a preliminary model that is trained using existing real world COVID-19 data.

**Methods:** We have utilized the computerized data of Maccabi Healthcare Services a 2.3 million member state-mandated health organization in Israel. The age and sex matched matrix used for training the XGBoost ILI-based model included, circa 690,000 rows and 900 features. The available dataset for COVID-based model included a total 2137 SARS-CoV-2 positive individuals who were either not hospitalized (n = 1658), or hospitalized and marked as mild (n = 332), or as having moderate (n = 83) or severe (n = 64) complications.

**Findings:** The AUC of our models and the priors on the 2137 COVID-19 patients for predicting moderate and severe complications as cases and all other as controls, the AUC for the ILI-based model was 0.852[0.824–0.879] for the COVID19-based model – 0.872[0.847–0.879].

**Interpretation:** These models can effectively identify patients at high-risk for complication, thus allowing optimization of resources and more focused follow up and early triage these patients if once symptoms worsen.

**Funding:** There was no funding for this study

**Research in context:** *Evidence before this study:* We have search PubMed for coronavirus[MeSH Major Topic] AND the following MeSH terms: risk score, predictive analytics, algorithm, predictive analytics. Only few studies were found on predictive analytics for developing COVID19 complications using real-world data. Many of the relevant works were based on self-reported information and are therefore difficult to implement at large scale and without patient or physician participation.

*Added value of this study:* We have described two models for assessing risk of COVID-19 complications and mortality, based on EMR data. One model was derived by combining a machine-learning model for influenza-complications with epidemiological data for age and sex dependent mortality rates due to COVID-19. The other was directly derived from initial COVID-19 complications data.

*Implications of all the available evidence:* The developed models may effectively identify patients at high-risk for developing COVID19 complications. Implementing such models into operational data systems may support COVID-19 care workflows and assist in triaging patients.

## Introduction

Since January 2020, the COVID-19 pandemic has become a global emergency. Healthcare organizations and governments, worldwide, are strained due to shortage of resources and the need to make timely decisions based on very little reliable data. These decisions include – who to test, how to treat positive cases, how to manage social distancing and reach-out to population at risk, contact tracing, and more. Many of these decisions could benefit from decision support tools based on EMR and additional data sources, such as geospatial information. Unfortunately, accurate data-driven tools are still difficult to develop due to the limited availability of COVID-19 patients’ data with historical EMR records. Many of the relevant works^1–3^ describe risk factors and the tools already developed^4,5^are based on self-reported information and are therefore difficult to implement at large scale and without patient or physician participation. Here, we describe two approaches and tools to assess the individual risk of developing COVID-19 complications based on medical records: a model developed by combining a machine-learning approach for influenza-like illness (ILI) to be used as a proxy model for COVID-19 and a second model using data on COVID-19 patients.

## Methods

### Settings

The models were trained using data from Maccabi Health Service (MHS) – a large Israeli HMO with a central EMR database containing longitudinal data for 2 million active individuals each year between 2010 and 2018. The data included full EMR information – demographics (e.g. age and sex), behavioral info (smoking status), vital signs, lab test results, diagnoses and procedures (using the *International Classification of Diseases* 9^th^ version), medication prescriptions and purchases, and hospital admissions (dates and departments only).

### Analytic approach

Since the number of in MHS members who are positive for SARS-CoV-2 is relatively low, and the data available is biased due to the current limitations of tests and challenges of data collection and curation, we have therefore chosen to test two complimentary approaches. First, we use a proxy model that we derived for identifying patients with high risk of developing complications due to influenza and apply some required adjustments. Although Influenza and COVID-19 are clearly very different diseases^6^, it is already apparent that both diseases have common risk factors for developing complications. However, the initial epidemiological data for COVID-19 [China CFR, NYC CFR] already show some major differences between the two diseases – primarily in the effect of increased age on the risk of complications (which seems much stronger for COVID-19) and the much higher risk among men for COVID-19 complications and mortality, a trend less evident in Influenza (Another difference is seasonality – which is clear for influenza and less evident for COVID-19). Following these differences, we modified the ILI-based model and forced it to ignore age and sex as risk factors, and then used Bayesian correction to add these risk factors using external priors.

For the training COVID-19ased model, we used information on SARS-CoV-2 positive individuals aged 19 or above within the MHS population, as well as information regarding hospitalization and in-hospital complications. As an initial prior we used the information based on COVID-19 mortality available from China [https://www.worldometers.info/coronavirus/coronavirus-age-sex-demographics/] as proxy for complications probabilities (appendix table 1). Fatality rate by sex is given in appendix table 2. Due to the over-representation of women among the elderly, we had to replace the 1:1.65 ratio of female-to-male risk with a higher 1:2 ratio per age group, as shown in appendix table 3.

**Table 1.** Excess risk of underlying health conditions compared to information from the CDC.

|  | CDC |  |  |  | MHS |  |  |  |
| --- | --- | --- | --- | --- | --- | --- | --- | --- |
| **Underlying condition** | **n** | **%** | **P(ICU)** | **Lift** | **n** | **%** | **ILI-based Model Lift** | **COVID19-model Lift** |
| None | 4470 | 62.4 | 0.02 | 1.00 | 573546 | 36.65 | 1.00 | 1.00 |
| One or above | 2692 | 37.6 | 0.13 | 6.00 | 1003410 | 63.35 | 4.41 | 2.79 |
| Diabetes* | 784 | 10.9 | 0.19 | 8.52 | 366068 | 22.96 | 7.45 | 4.74 |
| Chronic Lung disease | 656 | 9.2 | 0.14 | 6.47 | 539433 | 34.03 | 4.27 | 2.42 |
| Cardiovascular disease | 647 | 9 | 0.20 | 9.21 | 135822 | 8.55 | 12.53 | 6.82 |
| Immunosuppression | 264 | 3.7 | 0.16 | 7.01 | 115850 | 7.30 | 4.34 | 2.29 |
| Chronic kidney disease | 213 | 3 | 0.26 | 11.87 | 44097 | 2.77 | 17.84 | 8.89 |
| Pregnancy | 143 | 2 | 0.03 | 1.26 | 39734 | 2.55 | 0.16 | 0.35 |
| Neurologic disorder | 52 | 0.7 | 0.13 | 6.08 | 332967 | 21.00 | 6.19 | 3.36 |
| Chronic liver disease | 41 | 0.6 | 0.17 | 7.71 | 44510 | 2.79 | 8.13 | 4.29 |

**Table 2.** Confusion matrix of COVID-19 complications and the model predictions.

|  | COVID19-based Model |  |  | ILI-based Model |  |  |
| --- | --- | --- | --- | --- | --- | --- |
|  | Low Risk | Intermediate Risk | High Risk | Low Risk | Intermediate Risk | High Risk |
| No hospitalization | 1349 | 196 | 113 | 1369 | 187 | 102 |
| Mild complications | 170 | 85 | 77 | 181 | 70 | 81 |
| Moderate complications | 18 | 18 | 47 | 18 | 32 | 33 |
| Severe complications | 8 | 16 | 40 | 10 | 20 | 34 |

### Model Derivation

A detailed description of our approach to developing a model based on EMR data is given elsewhere^7,8^. For training the ILI-based model, a training set of all MHS members at September 1^st^ of every calendar year who were not vaccinated during the following flu-season. We marked them as cases if they were diagnosed with ILI followed by complications (death, hospitalization in internal ward, or severe illness, e.g. pneumonia – see appendix for list of ICD-9 codes) within 3 months, and controls if otherwise. Bins were matched for age (5yegendar groups) and sex. In addition, we matched for calendar year to avoid biases due to change in collection, registration, or healthcare policy over the period.

Given the matched set, we generated a large matrix of features per each sample (a sample corresponds to an individual per each relevant year) and applied a process of univariant age-and-sex conditioned feature selection on this matrix, and then trained and recalibrated XGBoost model^9^ using isotonic regression. To combine the prediction of the calibrated model with age and sex priors for complications, we used the following formula –

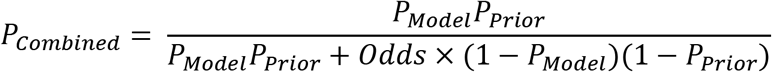

Where odds is the overall odds of the model’s predictions.

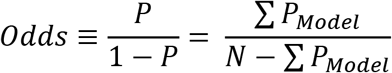

See the appendix for derivation of the formula.

### COVID-19-based model

We used the definitions of the Israel Ministry of Health for COVID19 complications: moderate (defined as pneumonia, with one of the following: respiratory rate above 30 breaths per minute, Respiratory distress, or oxygen saturation below 90%) or severe (pneumonia accompanied by sepsis, shock, ARDS or death). We then created a vector that of features per each individual, including risk factors and underlying conditions (see appendix). We used XGBoost on the features matrix to learn a COVID-19 complications predictor based on these features.

### Performance evaluation

Given that real world data on COVID-19 are currently limited, it is difficult to evaluate the performance of our models. We report here several methods we have used to estimate the value of the models.

1. For the COVID19-based model, we report the performance (AUC) in predicting influenza complications as an initial indication of the value of the models
2. We examined the excess risk of underlying health conditions, compared to information from the CDC [https://www.cdc.gov/mmwr/volumes/69/wr/mm6913e2.htm#F1_down]. For CDC information, we took the proportion of individuals admitted to ICU given various comorbidities; for our models, we used the mean prediction over individuals with the corresponding ICD-9 codes. Evaluating performance of the model on initial COVID-19 complications records. For the model directly derived on COVID-19 data, we used cross-validation for performance evaluation. Lift was evaluated by calculating the average prediction over the population with the underlying conditions, and comparing to the average prediction over a reference population

## Results

### Influenza-Complications Model

The age and sex matched matrix used for training the XGBoost model included, after feature selection, about 690,000 rows and 900 features (compared to about 790,000 rows and 1584 features for the non-matched model). The top 10 important features are given in an appendix.

### COVID-19-based model

The available dataset included a total 2137 SARS-CoV-2 positive individuals who were either not hospitalized (n = 1658), or hospitalized and marked as mild (n = 332), or as having moderate (n = 83) or severe (n = 64) complications. Individuals who were hospitalized but not assigned severity level were excluded. All individuals were linked to their MHS medical record in order to generate the features matrix

### Performance Evaluation

The AUC of the full (non-matched) model for predicting influenza-complication was 0.744, the matched model AUC was 0.726. After adjusting for the age and sex priors, the AUC for predicting influenza-complications deteriorates to 0.688.

In comparing our results to CDC data (table 1), we can see that, although the prevalence of various conditions is quite different between MHS and CDC data, lifts seem similar and correlated, both for the ILI-based and the COVID19-based models, with the exception of pregnancy.

We defined three groups according to the model’s prediction – high risk (top 10%), intermediate (next 15%) and low (bottom 75%).

The confusion matrix of our grouping and the MHS status is given in table 2. The overall distribution of SARS-CoV-2 positives over the three groups is as expected for random infection (ILI-based model: 74% for the low risk, 14% for the intermediate risk, and 1% for the high risk; 72%, 15% and 13% respectively for the COVID19-based model). The hospitalized population is enriched in high and intermediate risk groups (56% of 479 for the ILI-based model, 59% for the COVID19-based model). This sensitivity is even higher for the moderate (78% of 83 for both models) and severe cases (84% and 88% of 64). We also compared the performance of our score to using only the priors – the sensitivity of the severe and moderate cases in the top 25% (high and intermediate risk) is 81.0% [78.0–84.0] for the ILI-based model and 82.379.3%,85.3% the COVID19-based model, and 76.9% [73.5%-80.5%] for the priors information only. The difference in sensitivity is mainly due to younger individuals, aged 57 and less.

We also checked the AUC of our models and the priors on the 2137 SARS-CoV2 positives, marking moderate and severe complications as cases and all other as controls. The AUC for the ILI-based model was 0.852[0.824–0.879] for the COVID-19-based model –0.872[0.847–0.879] and 0.860[0.831–0.885] for using the priors.

The priors model is less continuous than our models. To demonstrate our models’ added value, we noted that 390 SARS-CoV2 positives patients receive the highest prior. Within this population the AUC for the ILI-based model was 0.61 [0.54,0.68] and 0.68 [0.62,0.73] for the COVID-19 based model

In comparing the two analytical models, BMI had a much larger effect in the COVID19-based model predictions. For example, the average risk-ratio for BMI 27kg/m^2^ as compared to and 22kg/m^2^, is 4.2-fold in the COVID19 model, compared to 2.3 for the ILI-based model. Other risk factors and underlying health conditions show lower contributions (by ∼30–40%) to the risk in the COVID19-based model, compared to the ILI-based model.

## Discussion

We have described two approaches for assessing risk of COVID-19 complications and mortality, based on EMR data. One model was derived by combining a machine-learning model for influenza-complications with epidemiological data for age and sex dependent mortality rates due to COVID-19. The other was directly derived from initial COVID-19 complications data.

Such models have many potential applications during the COVID-19 epidemics, prioritization of tests and antibody testing, follow-up on patients with the disease, decision on hospitalization, reach-out for population at risk when social-distancing and restrictions are gradually lifted, and in possible future outbreaks of the disease, and, hopefully in the near future, prioritization of vaccination. Both approaches have many weaknesses, due to the speedy and urgent manner of their derivations.

Performance evaluation is indicative, at best, of the true performance of the models. A better model will surely be derived once more reliable COVID-19 real world data will be available. However, we believe that currently such models can be of great use for health systems and public health entities coping with pandemic. Although performance of the COVID19-based model seems better than the ILI-based model, it is reasonable to suspect due to the small size of the dataset that the latter model is too specific to the MHS and less generalizable compared to the ILI-based model.

The AUC of the ILI-based model on the subset of SARS-CoV-2 positives is the same as the priors only. However, we note a couple of points – first, the significant difference in performance when considering all population, as well as some manual curation of the dataset suggest a possible bias toward older individuals in the definition of COVID-19 complications and SARS-CoV2 positives. Second, even though the AUC is similar, the ILI-based model can identify younger populations at risk of complications, which the priors-only model, of course, cannot.

In comparing the two models it is interesting to note that the effect of BMI on the risk for COVID-19 complications seems much higher than the risk for influenza complications. This suggests future work of further adjusting the more robust ILI-based –model by inserting exterior priors for BMI as well.

In comparing our results to CDC data similar lifts both for the ILI-based and the COVID19-based models, with the exception of pregnancy that was associated with low risk in our model, compared to slightly elevated risk in the CDC data (though based on very few cases). This might be due differences in age and sex distribution between the US and Israel populations pregnancy. For all underlying conditions, the COVID19-based model showed lower lift compared to the ILI-based model

Despite the model’s inherent weaknesses, and due to the clear and urgent needs, the ILI-based model was integrated at MHS to support two COVID-19 care workflows. First use is triaging testing. With limited resources available for outpatient testing, there is a need to prioritize testing to those individuals at highest risk of complications and mortality from COVID-19. The second use is for outpatient virtual management and triage. MHS established a virtual COVID-19 management center, occupied by primary care physicians and nurses. The medical staff are the first to contact confirmed COVID-19 patients, question them and decide on the appropriate treatment facility based on their symptoms and overall medical assessment. Patients can be hospitalized, sent to a special COVID-19 care facility, or stay at home. Nurses are then following-up on those patients at homecare to continuously assess their condition. A flag was added to patients estimated to be at high-risk, thus allowing optimization of resources and more focused virtual follow up and also helping clinicians to triage these patients if their symptoms worsen.

## Data Availability

Data referred to the study are not avaliable due to Israel privacy regulations

## Appendix

**Appendix Table 1.** Case fatality rate by age for the 2019–2020 Wuhan COVID-19 outbreak (https://www.worldometers.info/coronavirus/coronavirus-age-sex-demographics)

| Age group | Rate (%) |
| --- | --- |
| 19-30 | 0.1% |
| 30-39 | 0.1% |
| 40-49 | 0.4% |
| 50-59 | 1.3% |
| 60-69 | 3.6% |
| 70-79 | 8.0% |
| 80+ | 14.8% |

**Appendix Table 2.**
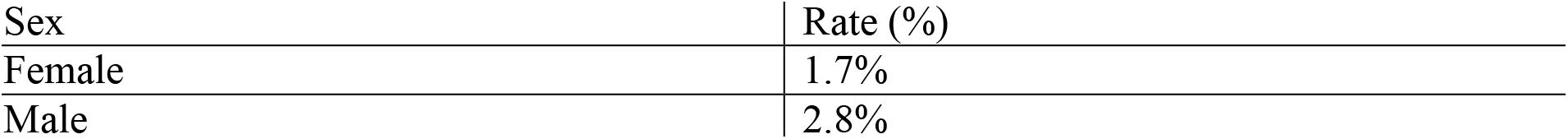
Case fatality rate by sex for the 2019–2020 Wuhan COVID-19 outbreak (https://www.worldometers.info/coronavirus/coronavirus-age-sex-demographics)

| Sex | Rate (%) |
| --- | --- |
| Female | 1.7% |
| Male | 2.8% |

**Appendix table 3.**
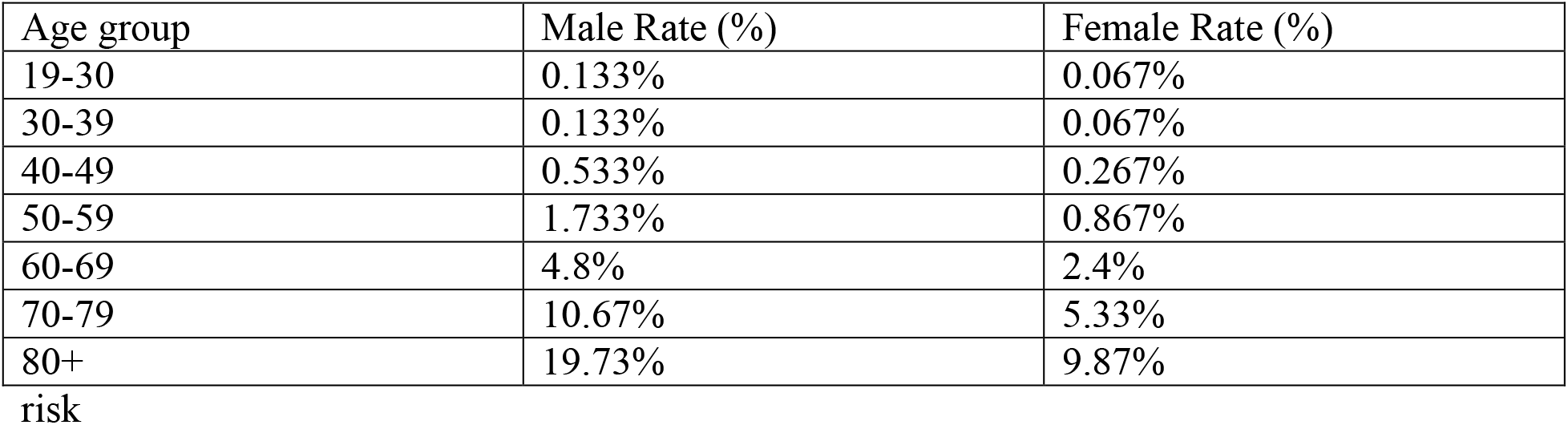
Age and sex dependent CFR used for Bayesian adjustment of COVID-19 complications

| Age group | Male Rate (%) | Female Rate (%) |
| --- | --- | --- |
| 19-30 | 0.133% | 0.067% |
| 30-39 | 0.133% | 0.067% |
| 40-49 | 0.533% | 0.267% |
| 50-59 | 1.733% | 0.867% |
| 60-69 | 4.8% | 2.4% |
| 70-79 | 10.67% | 5.33% |
| 80+ | 19.73% | 9.87% |

risk

## Appendix A Bayesian adjust

We learn a model for flu complications on an age-matched training set, and evaluate the following probabilities – *P_case_* = *P*(*Score* = *S*|*flu* + *Comp*) and *P_ctrl_* = *P*(*Score* = 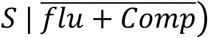;

We then use the following Bayesian argument –

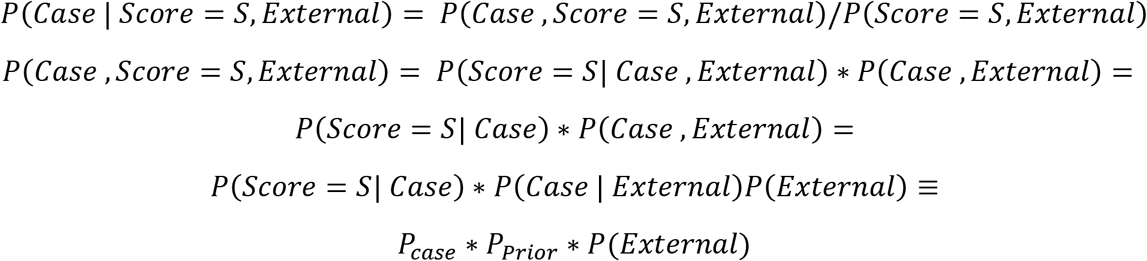

Using (*Score* = *S,External* = *P*(*Case, Score* = *S,External*) + *P*(*Ctrl, Score = S,External*), and applying the same argument to *P*(*Ctrl, Score = S,External*) we end with:

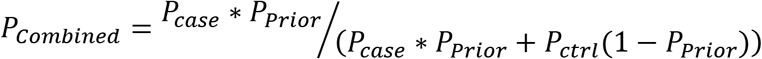

We now replace *P_case_* and *P_ctrl_* with *P_Model_* according to another application of Bayes rule,

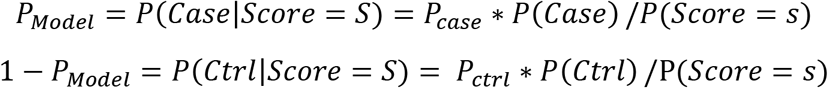

and get the formula in the paper.

## Appendix B Top features for the age and sex matched influenza complications model

- History of ICD9:466 Acute bronchitis and bronchiolitis
- History of Drug.ATC_R01: Nasal Preparations
- History of Drug.ATC_J01F: Macrolides, lincosamides and streptogramins
- History of Drug.ATC_J: Antiinfectives for systemic use
- History of ICD9:460–519: Diseases of The Respiratory System
- History of Drug.ATC_R:Respiratory system
- History of ICD:460–466: Acute Respiratory Infections
- History of Drug.ATC_R03: Drugs for obstructive airway diseases
- History of Drug.ATC_R01B: Nasal decongestants for systemic use
- History of Flu Complications

## Appendix C ICD-9 codes used for complications in the influenza-complications model

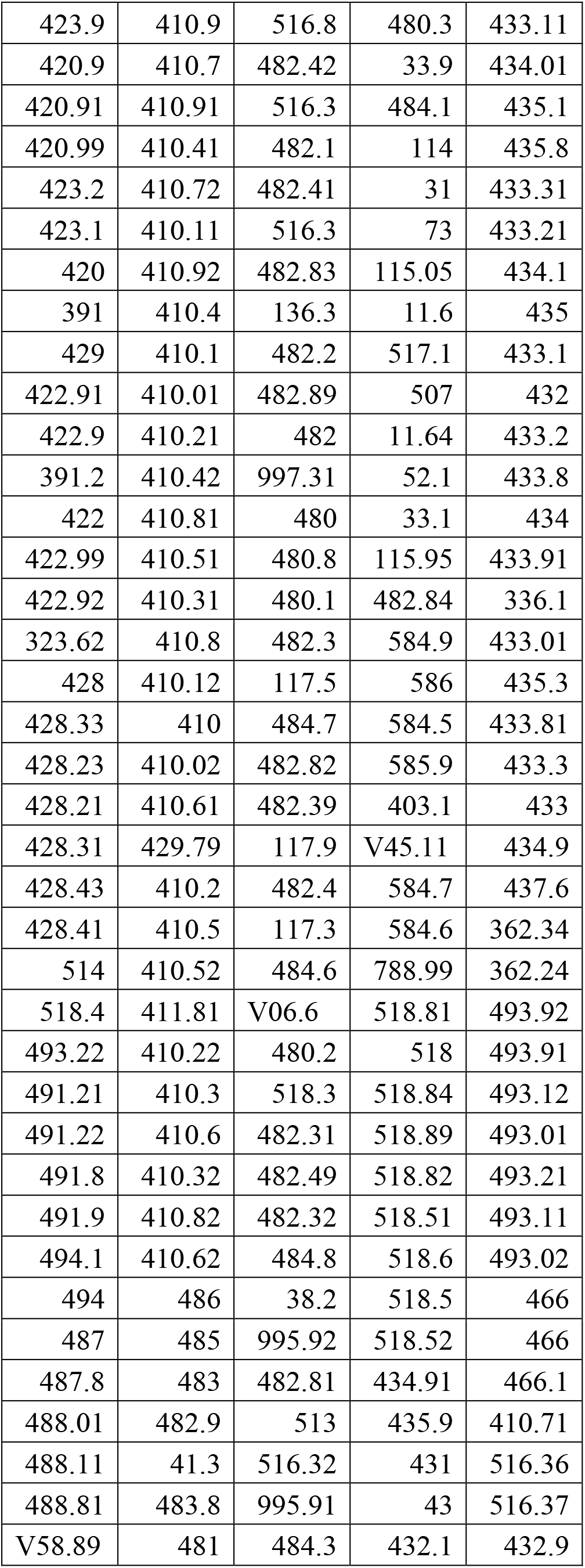

## Appendix D Features used in the COVID19-based model

- Age
- Sex
- BMI
- Indication of:
q
- Asthma
- Chronic Heart Disease
- Chronic Respiratory Disease
- Malignancy
- Diabetes Mellitus
- Immuno-suppression
- Chronic Kidney Disease
- Chronic Liver Disease
- Neurological Disorders
- Pregnancy
- COPD
- Indication of prescription of

- Asthma drugs
- Immuno-suppressing drugs
- ACE inhibitors
- Counts of

- Hospital admissions
- Recorded cases influenza
- Influenza complications.

